# Evaluating the performance and potential bias of predictive models for detection of transthyretin cardiac amyloidosis

**DOI:** 10.1101/2024.10.09.24315202

**Authors:** Jonathan Hourmozdi, Nicholas Easton, Simon Benigeri, James D. Thomas, Akhil Narang, David Ouyang, Grant Duffy, Ross Upton, Will Hawkes, Ashley Akerman, Ike Okwuosa, Adrienne Kline, Abel N. Kho, Yuan Luo, Sanjiv J. Shah, Faraz S. Ahmad

## Abstract

**Background:** Delays in the diagnosis of transthyretin amyloid cardiomyopathy (ATTR-CM) contribute to the significant morbidity of the condition, especially in the era of disease-modifying therapies. Screening for ATTR-CM with AI and other algorithms may improve timely diagnosis, but these algorithms have not been directly compared.

**Objectives:** The aim of this study was to compare the performance of four algorithms for ATTR-CM detection in a heart failure population and assess the risk for harms due to model bias.

**Methods:** We identified patients in an integrated health system from 2010-2022 with ATTR-CM and age- and sex-matched them to controls with heart failure to target 5% prevalence. We compared the performance of a claims-based random forest model (Huda et al. model), a regression-based score (Mayo ATTR-CM), and two deep learning echo models (EchoNet-LVH and EchoGo^®^ Amyloidosis). We evaluated for bias using standard fairness metrics.

**Results:** The analytical cohort included 176 confirmed cases of ATTR-CM and 3192 control patients with 79.2% self-identified as White and 9.0% as Black. The Huda et al. model performed poorly (AUC 0.49). Both deep learning echo models had a higher AUC when compared to the Mayo ATTR-CM Score (EchoNet-LVH 0.88; EchoGo Amyloidosis 0.92; Mayo ATTR-CM Score 0.79; DeLong P<0.001 for both). Bias auditing met fairness criteria for *equal opportunity* among patients who identified as Black.

**Conclusions:** Deep learning, echo-based models to detect ATTR-CM demonstrated best overall discrimination when compared to two other models in external validation with low risk of harms due to racial bias.

## Introduction

Using artificial intelligence (AI) and other machine-learning algorithms for the early detection or correcting misdiagnosis of rare diseases represents a major area of research and development.^1^ This area of research is particularly important for patients with transthyretin amyloid cardiomyopathy (ATTR-CM), a highly morbid condition that is increasingly recognized as an important cause of heart failure (HF) and disproportionately impacts Black communities.^2^ While the true prevalence of cardiac amyloidosis is unknown, recent data suggest that it is much more common than previously thought, comprising up to 17% of HF with preserved ejection fraction cases.^3^ Despite early clues to diagnosis, delays in the diagnosis of ATTR-CM are extremely common, with 42% of cases diagnosed over 4 years after the development of cardiac symptoms and a median time from first hospitalization to diagnosis of 2 to 3 years.^4^ Even prior to the development of clinical HF symptoms, subclinical changes in conduction, structure, and function often can be observed in cardiac testing, including routine electrocardiography and echocardiography.^5–8^

The emergence of transthyretin stabilizers, which can halt disease progression and dramatically improve morbidity for patients with ATTR-CM, as well as a pipeline of other disease-modifying therapies, have galvanized efforts to improve early and accurate diagnosis of ATTR-CM, including the use of AI and other algorithms for screening.^9^ In order to better understand the potential implications of implementing algorithmic screening technologies for ATTR-CM in large health systems, we studied the performance of four models for the identification of cardiac amyloidosis in a retrospective cohort of patients treated across an integrated health system with 11 hospitals and over 200 sites. In addition to overall diagnostic performance, we assessed the performance of each model with respect to selected fairness metrics to identify any potential risks to currently disadvantaged groups owing to possible algorithmic bias.

## Methods

The data for this study will be made available by the corresponding author upon reasonable request and a data use agreement. The Northwestern University Institutional Review Board approved this study. This paper adheres to the TRIPOD-AI guidance for reporting of clinical prediction models (**See Supplemental Checklist**).^10^

### Data Source and Participants

To identify patients with ATTR-CM, we first searched the Northwestern Enterprise Data Warehouse (NMEDW) for patients with at least 1 ICD code for amyloidosis, a pyrophosphate (PYP) scan, or medication order for Tafamidis from 1/1/2010 to 12/31/2022. For these patients, we performed a review of their records to confirm diagnosis of ATTR-CM, defined as either positive endomyocardial biopsy or PYP scan with a final reading of “strongly suggestive” and absence of serum evidence of amyloid light chain (AL) amyloidosis or negative bone marrow biopsy result. For a subset of patients with potential ATTR-CM based on our screening criteria but without available definitive testing in the NMEDW, we used data from the Northwestern Medicine Cardiac Amyloidosis Program registry (August 2018 to December 2022), which included the results of endomyocardial biopsies or PYP scans performed outside of the Northwestern Medicine system.

We then created a matched cohort of HF controls with at least one encounter ICD code for HF over the study period. We excluded potential cases of amyloidosis from this matched cohort, defined as at least one ICD code for amyloidosis and did not have definitive negative testing available, specifically a negative endomyocardial biopsy result or a PYP scan with a final reading of “Not Suggestive.” All cases and controls were required to have a transthoracic echocardiogram (TTE) up to six months before or any time after definitive diagnosis by positive endomyocardial biopsy or PYP scan. The NMEDW query required that these TTEs also reported measurements for left ventricular ejection fraction (LVEF), posterior wall thickness, and relative wall thickness in a format available for automated data extraction and inclusion in the Mayo ATTR-CM Score model. If controls had multiple TTEs, the one nearest to the first HF diagnosis code was used for our model evaluation. We matched controls to cases by age, sex, and year that TTE was obtained using the PsmPy package in Python.^11^ Logistic regression was performed to generate propensity logits using PsmPy’s resampling method to account for class imbalance. Cases were then matched to controls in a 19:1 ratio using the nearest neighbor method without caliper, to target a prevalence of ATTR-CM of 5%, which was selected to mirror estimates in the HF population and reflect its potential clinical use as screening tool in a broad population.^11^ We then excluded patients in which any of the model’s predictions were not available due to missing data, image quality or other factors.

### Cardiac Amyloidosis Risk Models

We compared the performance of four risk models with different inputs and model architectures **(Table 1)**. The Huda et al. model^12^ was developed to predict risk of wild-type ATTR-CM using databases of claims data and was validated on claims data and EHR data from patients with ICD codes for amyloidosis (including for codes beyond the specific code for wild-type ATTR). The Huda et al. model uses tabular, ICD code data in a random forest model. The Mayo ATTR-CM Score was developed by Davies et al.^13^ and uses tabular data that includes demographics (age, sex), history of hypertension (based on ICD codes for this analysis), and TTE measurements (left ventricular ejection fraction, wall thickness). The Mayo ATTR-CM Score is an integer-based score created from a linear regression model. Duffy et al.^14^ developed the EchoNet-LVH model, which is a computer vision model that uses a convolutional neural network architecture. EchoNet-LVH generates a prediction for likelihood of cardiac amyloidosis (any type) from an echocardiogram study synthesizing information about wall thickness from the parasternal long axis view and additional information from the apical four chamber view. Lastly, EchoGo^®^ Amyloidosis is another deep-learning classifier developed by Ultromics which was trained to predict a diagnosis of cardiac amyloidosis from a single apical 4 chamber view video clip and received FDA clearance for commercial use in November 2024.^15^ EchoGo Amyloidosis produces a probability score (0-1) and an uncertainty score. If EchoGo Amyloidosis generated a probability score for multiple apical four chamber clips within an echo study, the least uncertain score was used.

**Table 1:** Description of the evaluated machine-learning models for detection of cardiac amyloidosis.

| Author (year) | Model Name | Model Architecture | Input Features | Outputs | Positive Case Threshold | Reported AUC<sup>a</sup> |
| --- | --- | --- | --- | --- | --- | --- |
| Huda et al. (2021) | N/A | Random Forest | Medical Claims | Probability (0-1) | $> 0.5$ | 0.80 |
| Davies et al. (2022) | Mayo ATTR-CM Score | Logistic Regression | Clinical Variables | Risk score (0-10) | $\geq 6$ | 0.84 |
| Duffy et al. (2022) | EchoNet-LVH | ResNet3D | Echo Images | Probability (0-1) | $\geq 0.8$ | 0.83 |
| Publication Pending; Validation data available on FDA website<sup>15</sup> | EchoGo Amyloidosis | Convolutional Neural Network | Echo Images | Probability (0-1); Label of Positive, Negative, or Uncertain because on probability and uncertainty scores | $\geq 0.06$ | Not reported |

<sup>a</sup>Values are from initial published performance in external validation cohorts.
AUC denotes area under the receiver-operating characteristic curve

### Statistical Analysis

We used standard descriptive statistics to summarize baseline variables with categorical variables reported as counts and proportions and continuous variables reported as mean and standard deviation (SD). Comparisons of demographic and baseline clinical features between cases and controls were performed using Welch’s T-test for continuous variables and Chi-squared testing for categorical variables. For each model, we report model discrimination with receiver operating characteristics reported as area under the curve (AUC). We also report the area under the precision-recall curve as average precision (AP), a measure which may be more informative in datasets with class imbalance.^16^ Where reported, 95% confidence intervals were calculated using the bootstrapping method. Pairwise comparisons of model discrimination with AUC between the Mayo ATTR-CM score were performed using the DeLong test.^17^ In addition, we report F1 score, accuracy, sensitivity, selectivity, positive predictive value, negative predictive value, and false negative rate for all models based on published thresholds or recommendations from the model developers as shown in Table 1.

### Sensitivity and Fairness and Bias Analyses

We conducted several sensitivity analyses to further examine the algorithmic performance under various clinical subpopulations and deployment decisions. For the clinical subgroup analyses, we first examined subgroups of patients with at least moderate left ventricular hypertrophy defined by posterior wall thickness measurements^18^ and those with reduced LVEF. To evaluate the tradeoff between sensitivity and specificity in the two AI echo models, we examined differences in performance at different thresholds. Lastly, we evaluated the performance of the two AI echo models in a larger sample in which we imputed the value of 0 for patients in whom the AI echo models were unable to generate a score.

In conducting our fairness and bias analysis, we compared three metrics that address fairness across a protected attribute. A protected attribute refers to a feature or characteristic of a patient that should not be used as the basis of decision-making and should be generally protected from discrimination. These attributes can be defined according to anti-discrimination law or in accordance with institutional values.^19^ We chose the protected attribute of self-identified race or ethnicity given the previously described existing disparities in diagnosis of ATTR-CM for racial and ethnic minorities in the United States. From previously published fairness criteria,^19^ we selected the following three to report in our primary analysis: *Demographic parity*, defined as the ratio of the proportion of patients predicted positive by the model between groups, *Predictive parity*, defined as the ratio of positive predictive value (or precision) of the model output between groups, and *Equal opportunity*, defined as the ratio of false negative error rate between groups. We chose to prioritize the equal opportunity fairness criteria in weighing the risk of harms due to the current imbalance in diagnostic error owing to under- or missed diagnosis in US Black individuals. All analyses were performed using the *Aequitas* package in Python 3.12.^20^ The default fairness threshold of 0.2 was used to determine whether significant unfairness was present for each metric based on the “80% rule” of disparate impact.^20^

## Results

### Demographic Characteristics

240 cases of confirmed ATTR-CM were identified by NMEDW query and matched to 4560 patients for a total of 4800 patients. There was a total of 1,432 patients (42 cases) in which either EchoNet-LVH, EchoGo Amyloidosis, or both did not generate predictions due to factors including insufficient clip length, low image quality, insufficient videos by view classification, low predictive confidence, or uncertainty. The exclusion of these patients resulted in a final analytic cohort size of 3368 patients containing 176 confirmed cases of ATTR-CM matched to 3192 controls (**Figure 1**). In this cohort, mean age was 78.8 years (SD 9.5 years), 20.9% were female, 79.1% self-identified as White, and 9.0% self-identified as Black. Complete demographic and baseline clinical characteristics for case and control cohorts are shown in **Table 2**. As compared to age and sex-matched controls with HF, the cohort of patients with ATTR-CM had fewer patients who identified as White (58.5% vs. 80.3%) and more left ventricular hypertrophy (LVH) as quantified by their baseline TTE (mean posterior wall thickness of 15.6 mm vs. 11.1 mm). The demographics and clinical characteristics observed in the subgroup of patients excluded due to missing or uncertain predictions are provided in **Supplemental Tables 1 and 2**.

**Figure 1:**
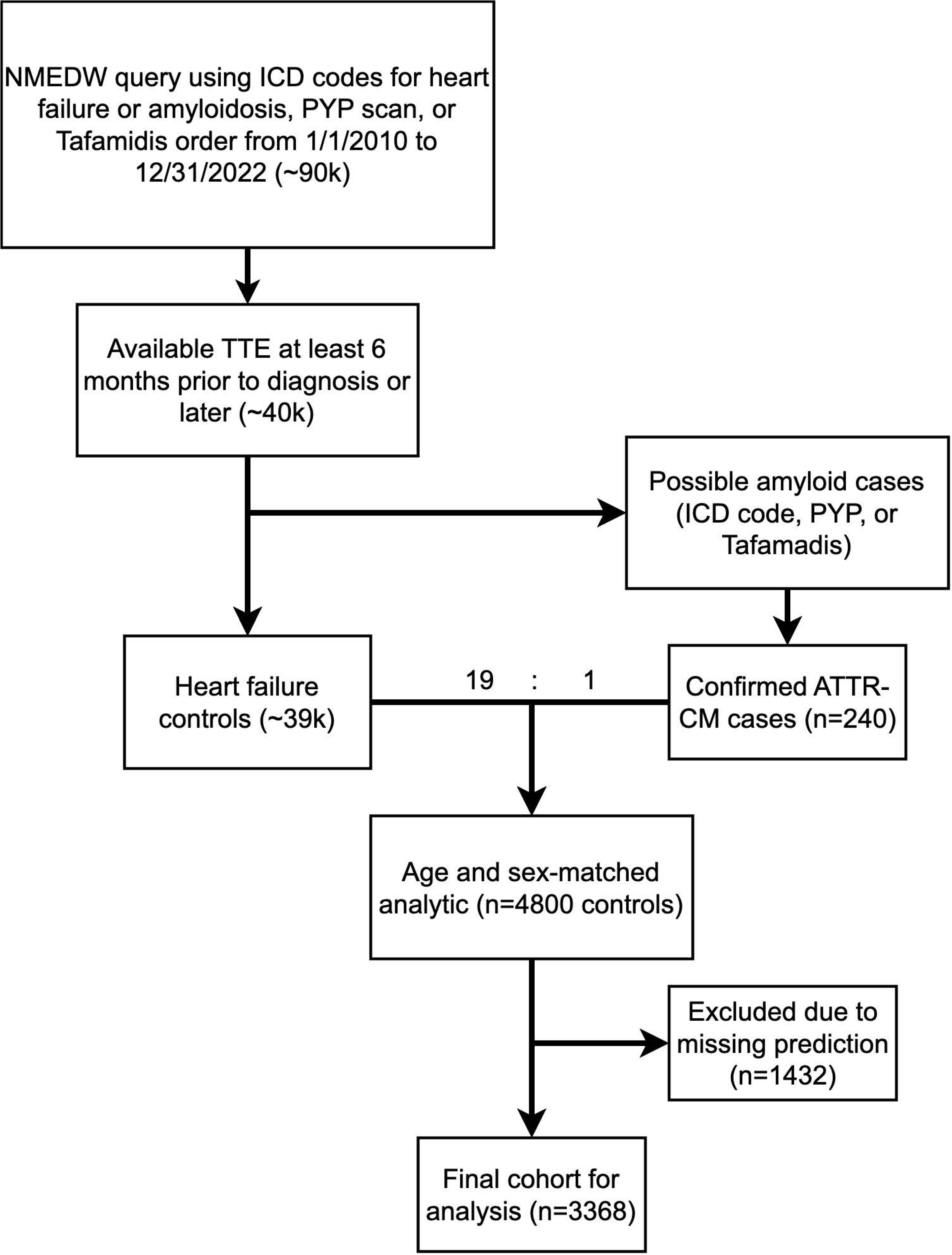
Flow diagram for analytic cohort creation. This figure shows the creation of the analytical dataset. NMEDW denotes Northwestern Medicine Enterprise Database Warehouse; ICD, international classification of diseases; TTE, transthoracic echocardiogram; PYP, pyrophosphate scan.

**Figure 2:**
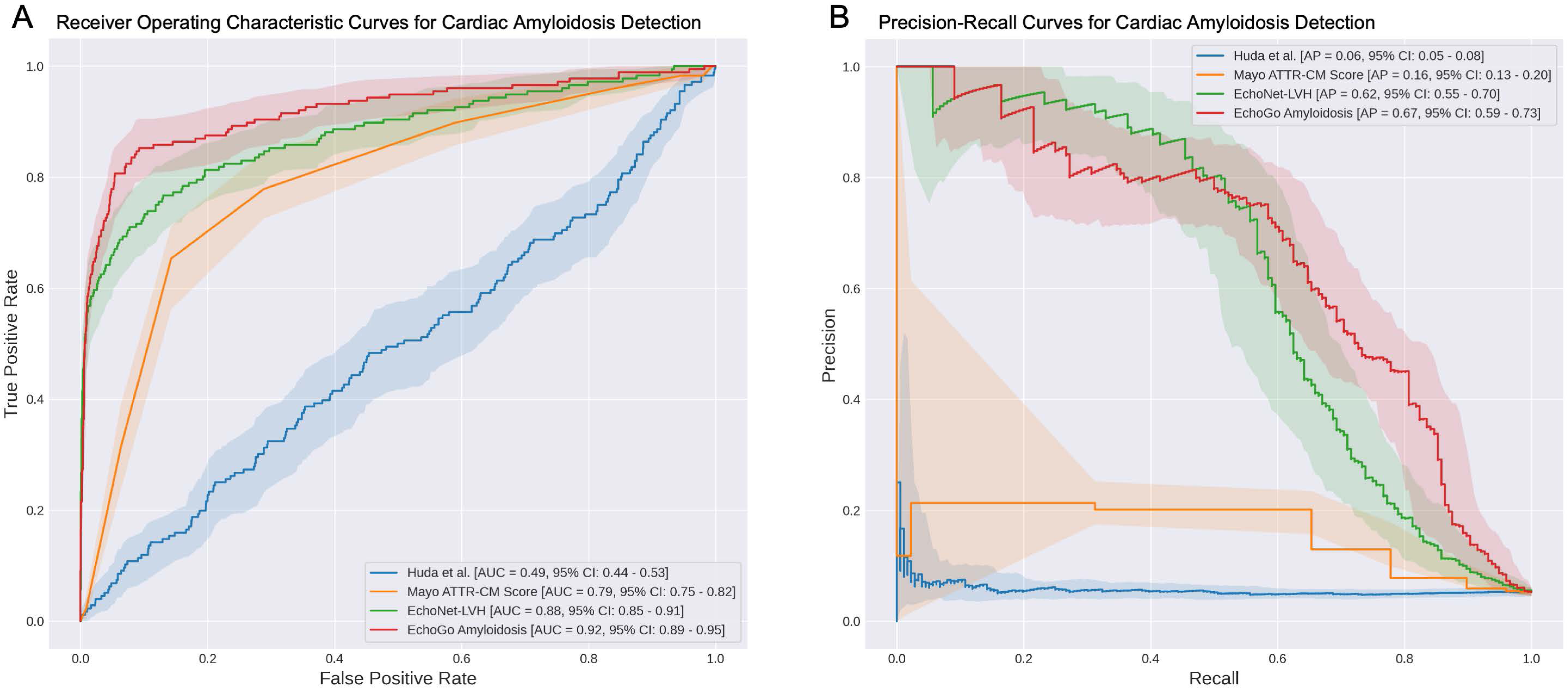
Comparison of model performance. Panel A shows receiver operator characteristic curves and Panel B shows the precision-recall curves (bottom) with bootstrap confidence intervals.

**Table 2:** Demographic characteristics of the validation cohort.

|  | ATTR-CM<br>(n=176) | HF Controls<br>(n=3192) | P-value | Overall<br>(n=3368) |
| --- | --- | --- | --- | --- |
| Age, mean (SD) | 78.6 (9.0) | 78.8 (9.5) | 0.805 | 78.8 (9.5) |
| Sex, n (%) |  |  | 0.466 |  |
| Female | 33 (18.8%) | 670 (21.0%) |  | 703 (20.9) |
| Male | 143 (81.2%) | 2522 (79.0%) |  | 2665 (79.1) |
| Race/Ethnicity, n (%) |  |  | < 0.0001 |  |
| Black | 59 (33.5) | 243 (7.6) |  | 302 (9.0) |
| Hispanic | < 10<sup>a</sup> | 127 (4.0%) |  | -<sup>a</sup> |
| White | 103 (58.5) | 2563 (80.3) |  | 2666 (79.2) |
| Other/Unknown | < 10<sup>a</sup> | 259 (8.1%) |  | -<sup>a</sup> |
| LVEF, n (%) |  |  | 0.142 |  |
| < 40% | 34 (19.3%) | 679 (21.2%) |  | 713 (21.2%) |
| 40% to < 50% | 35 (19.9%) | 451 (14.1%) |  | 486 (14.4%) |
| ≥ 50% | 107 (60.8%) | 2062 (64.6%) |  | 2169 (64.4%) |
| LVH by Posterior Wall Thickness<sup>b</sup> |  |  | < 0.0001 |  |
| Mean, (SD) | 15.6 (4.0) | 11.1 (2.2) |  | 11.3 (2.5) |
| Normal/Mild, n (%) | 56 (31.8%) | 2800 (87.8%) |  | 2856 (84.8%) |
| Moderate, n (%) | 57 (32.4%) | 333 (10.5%) |  | 390 (11.6%) |
| Severe, n (%) | 63 (35.8%) | 51 (1.6%) |  | 114 (3.4%) |
| LVH by Septal Wall Thickness<sup>b</sup> |  |  | < 0.0001 |  |
| Mean, (SD) | 15.9 (3.8) | 11.7 (2.9) |  | 12.0 (3.1) |
| Normal/Mild, n (%) | 42 (23.9%) | 2011 (63.0%) |  | 2045 (60.7%) |
| Moderate, n (%) | 48 (32.4%) | 380 (15.3%) |  | 428 (16.3%) |
| Severe, n (%) | 57 (38.5%) | 91 (3.7%) |  | 148 (5.6%) |
| Relative Wall Thickness |  |  | < 0.0001 |  |
| Mean (SD) | 0.78 (0.47) | 0.47 (0.15) |  | 0.49 (0.19) |
| > 0.57 (%) | 137 (77.8%) | 628 (19.7%) |  | 765 (22.7%) |
| Hypertension, n (%) | 144 (81.8%) | 2560 (80.2%) | 0.590 | 2704 (80.3%) |

<sup>a</sup> Cell sizes not reported due to small cell size<sup>b</sup> Sex-specific cutoffs were used from consensus recommendations<sup>18</sup>
ATTR-CM, transthyretin amyloid cardiomyopathy cases; HF, heart failure; LVEF, left ventricular ejection fraction; LVH, left ventricular hypertrophy; SD, standard deviation; IQR, interquartile range.

### Overall Model Performance and Comparison

In terms of overall performance, the Huda et al. model failed to achieve meaningful discriminatory performance with an overall AUC of 0.49 (95% CI, 0.44 to 0.54) and had relatively poor performance on all other metrics. Compared to the ATTR-CM Score (AUC of 0.79, 95% CI, 0.76 to 0.83), EchoNet-LVH (AUC 0.88, 95% CI, 0.85 to 0.91) and EchoGo Amyloidosis (AUC 0.92, 95% CI, 0.89 to 0.95) both outperformed the simpler model (both DeLong P<0.001). A complete comparison of performance metrics for all four models can be found in **Table 3**.

**Table 3:**
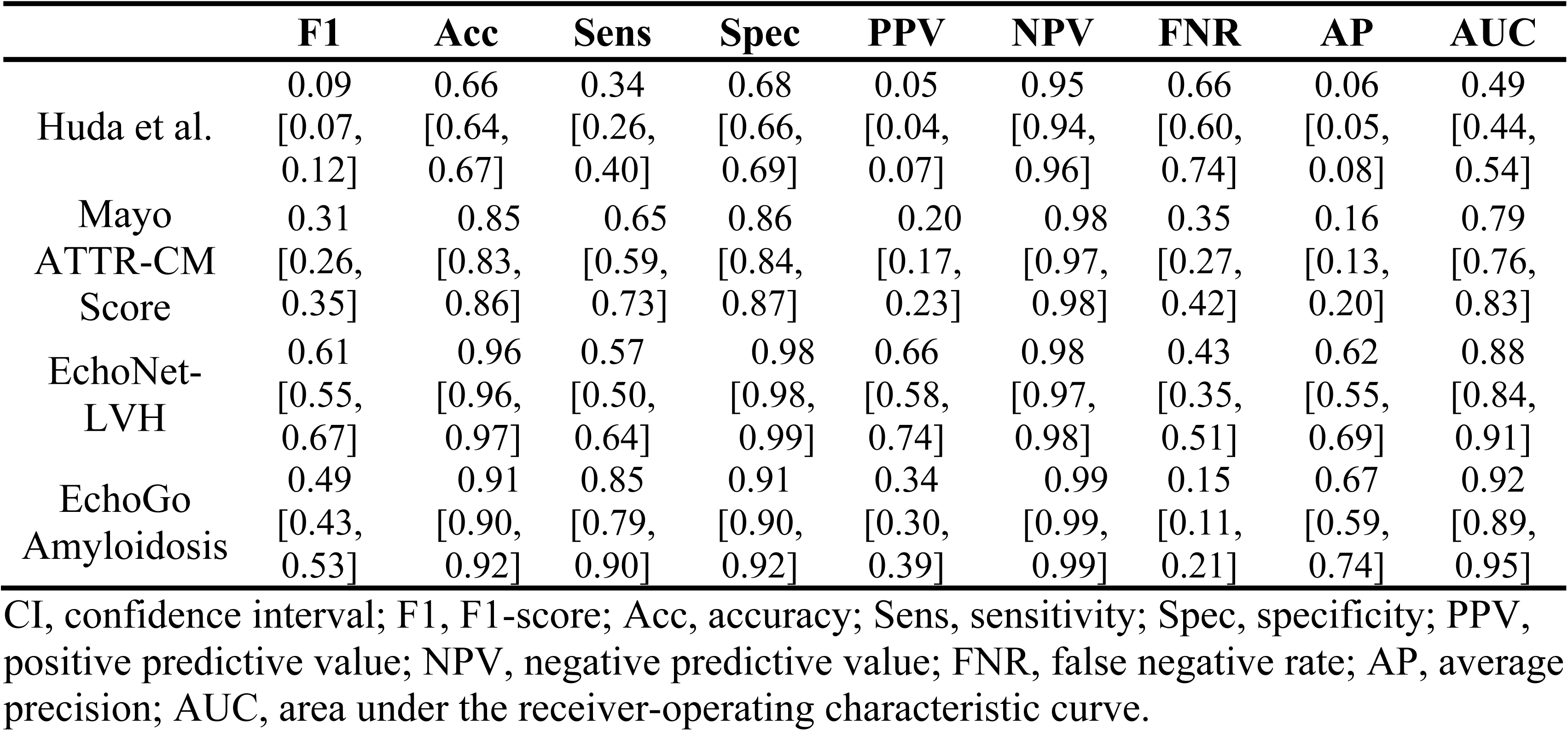
Overall performance metrics with 95% CI for the four compared models for 176 cases and 3192 controls at specified threshold by model developer.

From the initial analysis of the primary model performance metrics, it was apparent that the performance of the Huda et al. model was poor compared to the other two models. The AUC of 0.49 in the analytic cohort was much lower than the model’s performance in their initial external validation cohorts.^12^ To determine whether insufficient number of diagnosis codes was contributing to this decreased performance, we reassessed the Huda et al. model on the subset of patients in the overall analytic cohort for whom at least 50 encounters were available (106 cases and 774 controls). There was no significant improvement in the overall model performance (AUC 0.56, 95% CI, 0.50 to 0.62, De long P=0.084, **Supplemental Table 3**). Because of this, we removed the Huda et al. model from further analyses and focused on the other three models.

### Clinical Subgroup and Additional Sensitivity Analyses

In the subgroup of patients with at least moderate left ventricular hypertrophy (n=504), the prevalence of ATTR-CM was higher than in the overall cohort (23.8% vs 5.2%). Compared to the overall cohort, the AI echo models had modest differences in AUC, sensitivity, and specificity as demonstrated in **Table 4**. The lower AUC for the Mayo ATTR-CM score with non-overlapping confidence interval suggests worse performance in this subgroup. In the subgroup of patients with a reduced LVEF of lower than 40% (n=713, prevalence = 4.8%), overall similar performance was again noted for the two AI echo models. The higher AUC for the Mayo ATTR-CM score with non-overlapping confidence interval suggests an improvement in performance in this subgroup (**Table 5**). An analysis of the two AI echo models at different thresholds highlights the tradeoffs between sensitivity and specificity (**Supplemental Table 4**). The two AI echo models have overall similar performance when set at the same threshold, with some tradeoff between sensitivity and specificity. Lastly, we conducted a sensitivity analysis in those patients in whom the computer vision models were unable to generate predictions in which we imputed missing prediction as a negative (score=0) and found, as expected, both models had lower sensitivity, false negative rate, and AUC (**Supplemental Table 5**).

**Table 4:** Clinical subgroup analyses for the 504 patients (120 cases, 384 controls) with at least moderate left ventricular hypertrophy by echo (posterior wall thickness ≥1.4 for men, ≥ 1.3 for women).

|  | **Sensitivity**<br>**[95% CI]** | **Specificity**<br>**[95% CI]** | **AP [95%CI]** | **AUC**<br>**[95%CI]** |
| --- | --- | --- | --- | --- |
| **Mayo ATTR-CM Score** | 0.82<br>[0.74, 0.88] | 0.40<br>[0.35, 0.44] | 0.28<br>[0.24, 0.34] | 0.60<br>[0.55, 0.65] |
| **EchoNet-LVH** | 0.66<br>[0.57, 0.74] | 0.96<br>[0.94, 0.98] | 0.81<br>[0.74, 0.87] | 0.91<br>[0.88, 0.94] |
| **EchoGo Amyloidosis** | 0.92<br>[0.87, 0.96] | 0.85<br>[0.82, 0.88] | 0.83<br>[0.76, 0.89] | 0.93<br>[0.90, 0.95] |

CI, confidence interval; AP, average precision; AUC, area under the receiver-operating characteristic curve.

**Table 5:** Clinical subgroup analyses for Mayo ATTR-CM score, EchoNet-LVH, and EchoGo Amyloidosis in 713 patients (34 cases, 679 controls) with left ventricular ejection fraction < 40%.

|  | **Sensitivity**<br>**[95% CI]** | **Specificity**<br>**[95% CI]** | **AP**<br>**[95%CI]** | **AUC**<br>**[95%CI]** |
| --- | --- | --- | --- | --- |
| **Mayo ATTR-CM Score** | 0.71<br>[0.54, 0.86] | 0.92<br>[0.9, 0.94] | 0.24<br>[0.15, 0.36] | 0.83<br>[0.74, 0.90] |
| **EchoNet-LVH** | 0.59<br>[0.43, 0.77] | 0.98<br>[0.97, 0.99] | 0.65<br>[0.45, 0.79] | 0.88<br>[0.79, 0.95] |
| **EchoGo Amyloidosis** | 0.82<br>[0.69, 0.94] | 0.88<br>[0.85, 0.9] | 0.67<br>[0.49, 0.83] | 0.89<br>[0.79, 0.95] |

CI, confidence interval; AP, average precision; AUC, area under the receiver-operating characteristic curve.

### Fairness and Bias Audit

Prevalence of ATTR-CM was highest in the group of patients who self-identified as Black (19% compared to 3-5% in the other racial and ethnic groups). With respect to the protected attribute of race, all models met the fairness criteria for equal opportunity in patients who self-identified their race as Black **(Figure 3)**. A summary of model performance with respect to all chosen fairness criteria is provided in **Table 6**. A complete comparison of all absolute metrics by protected attribute can be found in **Supplemental Figures 1 to 3**.

**Figure 3:**
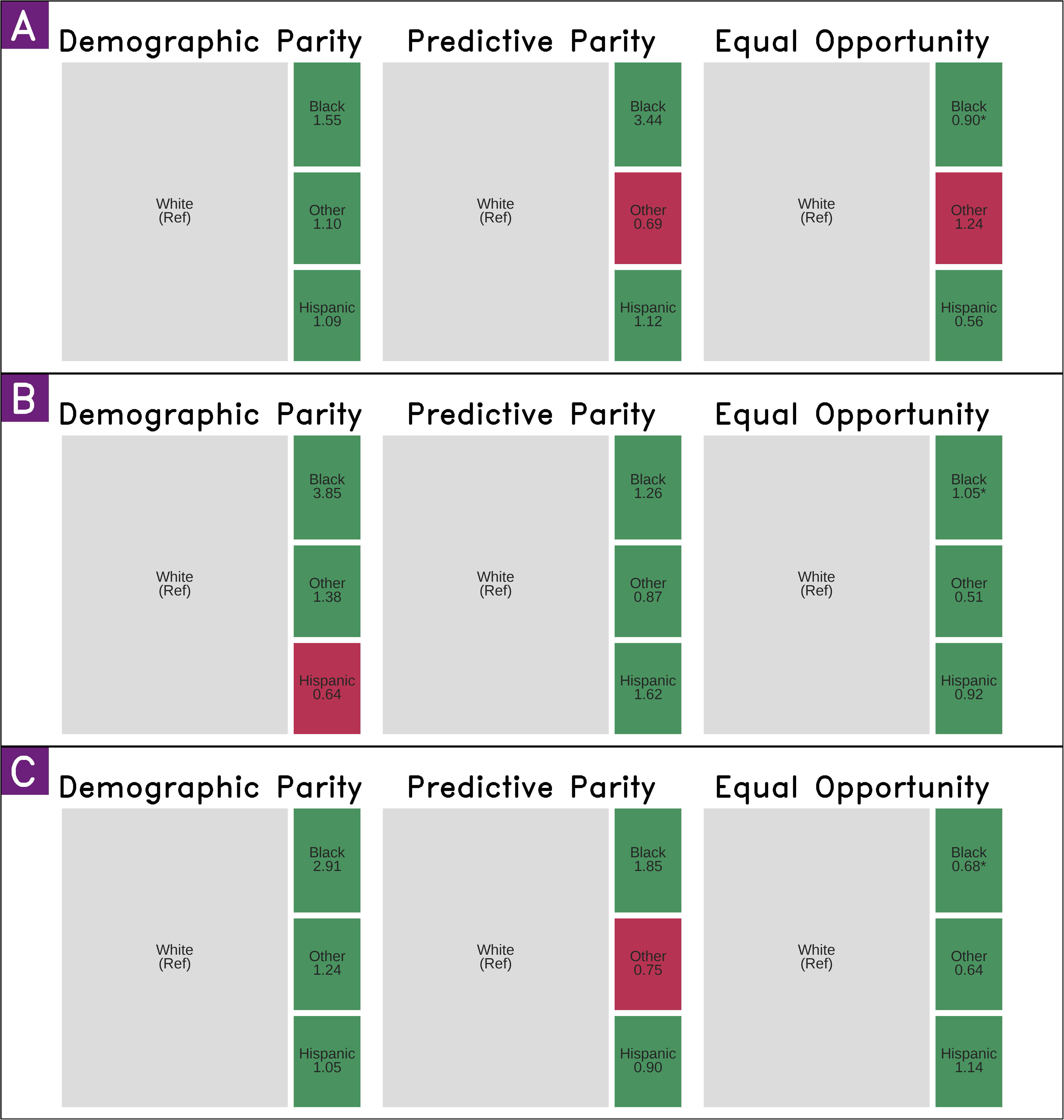
Parity metric treemaps for Mayo ATTR-CM Score model (A), EchoNet-LVH (B), and EchoGo Amyloidosis (C). Selected fairness metrics are expressed as a ratio of protected group: reference group. *Demographic parity* refers to positive prediction rate, *predictive parity* refers to positive predictive value (precision), and *equal opportunity* refers to false negative rate. Treemap squares are sized according to size of group in the cohort and colored red if results suggest disparity according to the “80% rule” of disparate impact.*\* denotes statistical significance of finding based on unadjusted alpha < 0.05*.

**Table 6:**
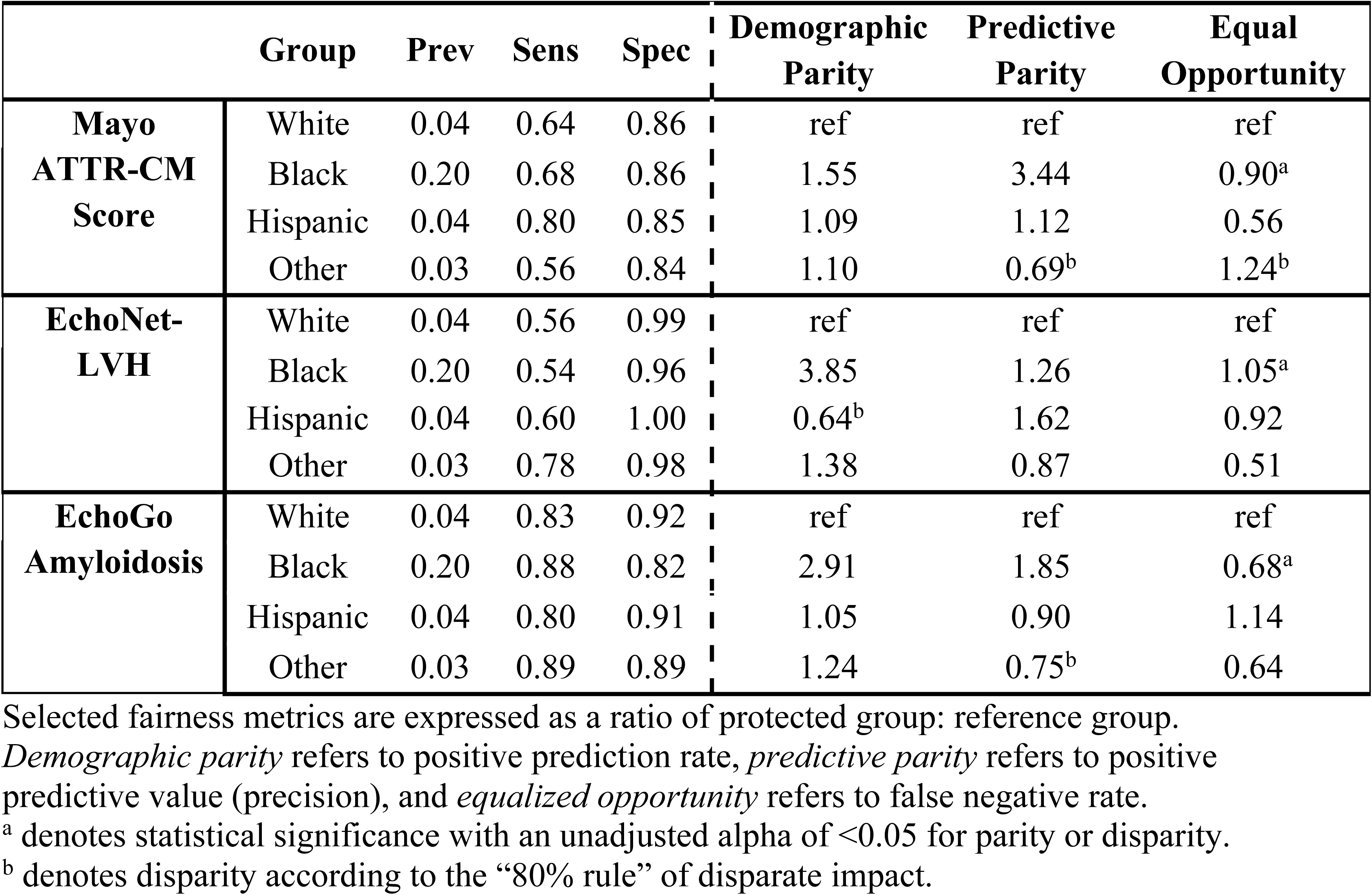
Summary of model performance across chosen fairness metrics stratified by protected attribute.

|  | Group | Prev | Sens | Spec | Demographic Parity | Predictive Parity | Equal Opportunity |
| --- | --- | --- | --- | --- | --- | --- | --- |
| **Mayo ATTR-CM Score** | White | 0.04 | 0.64 | 0.86 | ref | ref | ref |
|  | Black | 0.20 | 0.68 | 0.86 | 1.55 | 3.44 | 0.90<sup>a</sup> |
|  | Hispanic | 0.04 | 0.80 | 0.85 | 1.09 | 1.12 | 0.56 |
|  | Other | 0.03 | 0.56 | 0.84 | 1.10 | 0.69<sup>b</sup> | 1.24<sup>b</sup> |
| **EchoNet-LVH** | White | 0.04 | 0.56 | 0.99 | ref | ref | ref |
|  | Black | 0.20 | 0.54 | 0.96 | 3.85 | 1.26 | 1.05<sup>a</sup> |
|  | Hispanic | 0.04 | 0.60 | 1.00 | 0.64<sup>b</sup> | 1.62 | 0.92 |
|  | Other | 0.03 | 0.78 | 0.98 | 1.38 | 0.87 | 0.51 |
| **EchoGo Amyloidosis** | White | 0.04 | 0.83 | 0.92 | ref | ref | ref |
|  | Black | 0.20 | 0.88 | 0.82 | 2.91 | 1.85 | 0.68<sup>a</sup> |
|  | Hispanic | 0.04 | 0.80 | 0.91 | 1.05 | 0.90 | 1.14 |
|  | Other | 0.03 | 0.89 | 0.89 | 1.24 | 0.75<sup>b</sup> | 0.64 |

Selected fairness metrics are expressed as a ratio of protected group: reference group.
*Demographic parity* refers to positive prediction rate, *predictive parity* refers to positive predictive value (precision), and *equalized opportunity* refers to false negative rate.
<sup>a</sup> denotes statistical significance with an unadjusted alpha of <0.05 for parity or disparity.
<sup>b</sup> denotes disparity according to the “80% rule” of disparate impact.

## Discussion

In this study, we evaluated the potential screening performance of four available models to detect ATTR-CM in a large, integrated health system. We found that the Huda et al. model performed poorly compared to their previously published validation data and the other models.^12^ The Mayo ATTR-CM score had lower sensitivity and higher specificity than previously reported, likely reflecting the differences in the cohort in this study and the derivation and validation cohorts for the score.^13,21^ The AI echo models that we evaluated demonstrated similar performance in our validation cohort as compared to their initially reported results, including across a key subgroup as shown in the bias audit.^14,15^ To our knowledge, this is the first comparison of multiple algorithmic approaches to screening for cardiac amyloidosis using different data inputs (diagnosis codes, structured clinical data, and echo images) and models (regression, random forest, and convolutional neural networks).

In addition to external validation, our study allowed us to directly compare a regression-based, simple score against more complex, computer vision models in our patient population and provides realistic estimates for predictive values beyond AUC alone. The deep learning vision-based models including EchoNet-LVH and EchoGo Amyloidosis, both performed better in terms of positive predictive value compared to Mayo ATTR-CM Score (0.66 for EchoNet-LVH vs. 0.34 for EchoGo Amyloidosis vs. 0.20 for Mayo ATTR-CM Score). Although the Mayo ATTR-CM Score include selected echo measurements, this analysis illustrates that AI echo models are likely integrating multiple features for cardiac amyloidosis that are likely more granular that our standard measurements. This has important implications for implementation as a reduction in unnecessary downstream testing due to false positive predictions could help to offset the higher upfront costs of adopting and integrating a more complex screening model. However, the optimal threshold for each model that balances between sensitivity and specificity remains uncertain, as evident by the different choice in threshold by the two AI echo model developers. Identifying the optimal threshold will require additional, prospective evaluation and expert consensus on the appropriate population for screening, approaches for implementation, and targets for sensitivity, positive predictive value, and other clinically informative metrics.

Our study also highlighted the technical challenge of AI model portability. Both computer vision-based models did not provide predictions in approximately 15-20% of patients, leading to the exclusion of ATTR-CM cases in our cohort. Thus, the sensitivity and other related performance metrics for the algorithm may be lower in real-world practice than estimated by our primary analysis as shown in the Supplement. Similarly, the data required to run the Mayo ATTR-CM Score likely does not exist in a structured form in many EHR data repositories, thus limiting its ability to be run at scale in a health system. The poor performance of the Huda et al. model underscores the potential limitations of training models using diagnosis codes alone, as demonstrated in another cardiac amyloidosis study.^22^

### Risk of perpetuating bias

With any machine learning model, there is a risk that they can learn to exploit patterns in the training data that represent or perpetuate bias, including systemic and structural racism. These risks are exacerbated when there is underrepresentation of minority patient groups in training cohorts. In the United States, patients who identify as Black disproportionately bear the burden of missed and delayed diagnosis in ATTR-CM, leading to worse outcomes.^2^ This is despite a higher prevalence of ATTR-CM in many Black communities in the US; in our cohort, the relative prevalence of ATTR-CM was about 5-fold higher for Black patients compared to all other racial groups. Such context is instrumental in interpreting fairness metrics related to any screening AI model, as there is a risk not only of differential predictive performance, but also of a differential distribution of error rate. Our findings support that there is a low risk of harm due to algorithmic bias from adopting the use of any of these machine learning models for screening in our integrated health system. This study also provides an example of how to apply the recently published TRIPOD-AI reporting guidance and bias and fairness metrics to external validation studies.

### Study limitations

Importantly, a major limitation of our study is that we utilize retrospective data from a single large health system to perform our analyses. The prevalence in our cohort was artificially set to 5% based on realistic estimates of ATTR-CM prevalence in our HF patient population. Absolute measurements of predictive value vary depending on prevalence and may not generalize to all healthcare settings. However, comparisons of relative predictive value between models are more useful when considering realistic estimates of population prevalence.

Secondly, a limitation of our bias audit is that self-identified race and ethnicity represent a social construct. Concepts of race and ethnicity vary with respect to time and geography, and the results of our bias audit, while likely generalizable to our institution’s population of patients with HF, should not be taken to represent a blanket determination of fairness with respect to race in all settings. Instead, we encourage institutions and constituents to partner with health equity researchers and consider similar methodology when evaluating models for potential deployment across a health system. Lastly, the study population differed from the validation population used by the different models by factors such as age cutoffs, clinical factors, and control selection. However, our study sample reflects the potential group in which a health system may elect to deploy a population screening strategy for cardiac amyloidosis and thus is clinically informative.

## Conclusion

Machine learning techniques have shown promise when applied to imaging and EHR data to enhance the early detection or correct the misdiagnosis of diseases, such as cardiac amyloidosis. The use of such AI-assisted diagnostic tools in ATTR-CM could improve racial and ethnic disparities through the early identification and uptake of novel therapies in these patients. However, despite pioneering research and significant resources dedicated to this task, few algorithms have had a meaningful impact on the care of these patients in practice. The reasons for this are multi-factorial and include the lack of prospective, high quality evidence and consensus recommendations to support at scale implementation, barriers related to costs, infrastructure, and required personnel, and evolving regulatory guidance over the deployment and governance of AI and machine learning algorithms. Future work on algorithmic approaches for screening for cardiac amyloidosis should include evaluating the performance of using multiple models in serial or fusion approaches. However, ultimately, to evaluate the value of these algorithms, we need randomized, effectiveness-implementation hybrid trials that leverage health equity conceptual models and frameworks to study the performance of the algorithms and the implementation strategies for successful deployment across diverse populations.^23–25^

## Perspectives

### Competency in medical knowledge

Algorithmic approaches to screening for ATTR-CM vary in their performance in single health system with AI echo models having the best overall performance. Both simple, regression-based score or AI echo models performed similarly in a fairness and bias analysis in patients who identify as Black.

### Translational Outlook 1

Understanding the optimal thresholds for different algorithmic approaches and evaluating the impact of prospective deployment remain important questions.

### Translational Outlook 2

Evaluation of multiple algorithmic screening approaches, either in serial or in a fusion approach, may lead to higher performance and requires further evaluation.

## Supporting information

Supplemental Checklist_TRIPOD AI

## Data Availability

All data produced in the present study are available upon reasonable request to the authors and under the execution of a data use agreement.

## Abbreviation List

AI: Artificial intelligence
ATTR-CM: transthyretin amyloid cardiomyopathy
HF: heart failure
NMEDW: Northwestern Enterprise Data Warehouse
PYP: Pyrophosphate
TTE: transthoracic echocardiogram
AUC: area under the curve
AP: average precision
LVH: left ventricular hypertrophy
LVEF: left ventricular ejection fraction

---

**Central Illustration.**
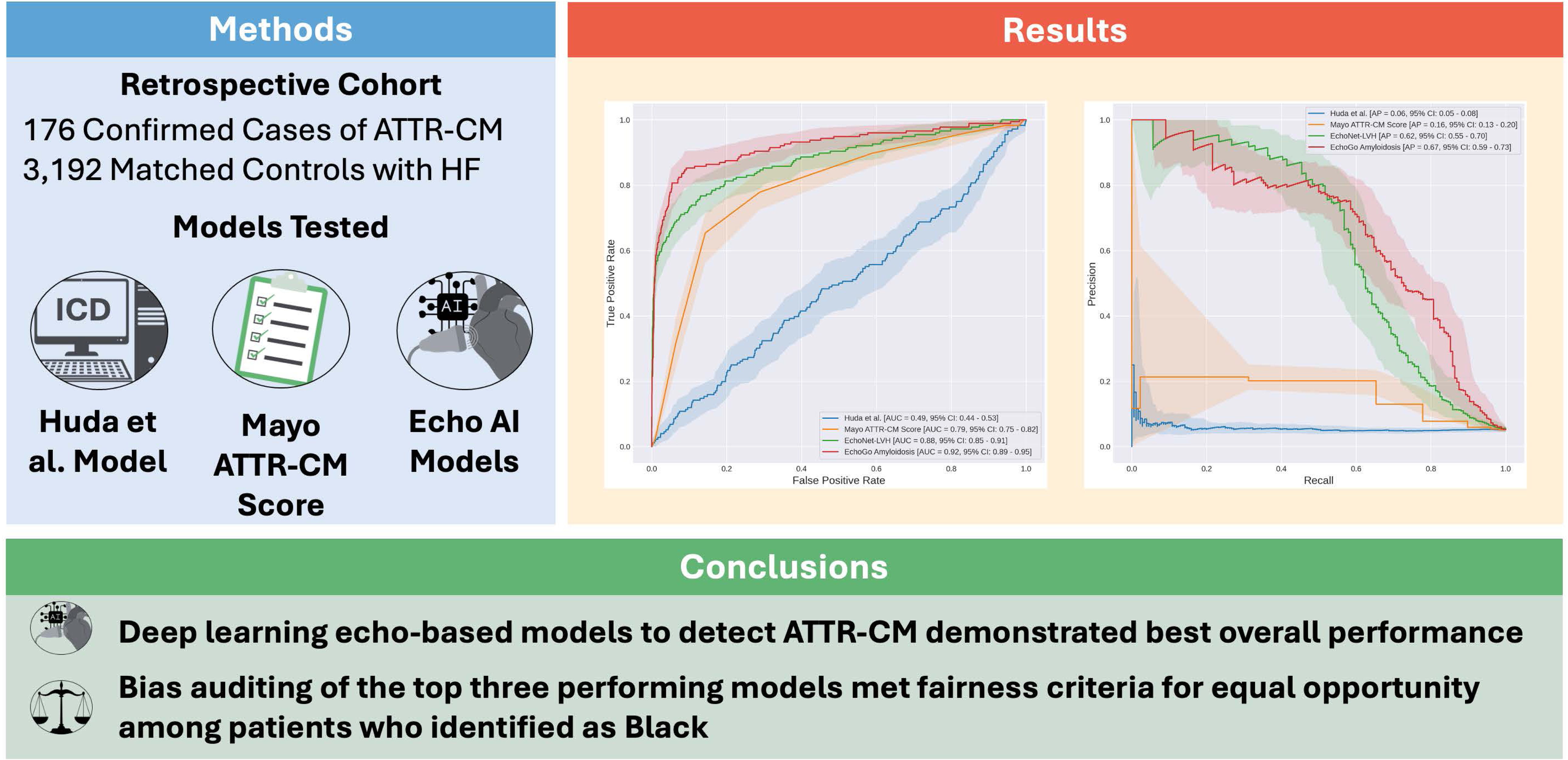
Evaluating the Performance and Potential Bias of Predictive Models for the Detection of Transthyretin Cardiac Amyloidosis. In a diverse cohort of 3416 patients with heart failure including 177 patients with transthyretin amyloid cardiomyopathy (ATTR-CM), deep-learning echocardiography models were best able to detect ATTR-CM compared to two other publicly available models and did not demonstrate any evidence of racial bias in a fairness audit. HF denotes heart failure; AUC, area under the receiver-operator characteristic curve; AP, average precision.

