## Supplemental Material for "Evaluating the performance and potential bias of predictive models for detection of transthyretin cardiac amyloidosis"

*Supplementary Appendix*

**Supplemental Figure 1. Absolute group metrics for Mayo ATTR-CM Score by race/ethnicity and sex.**
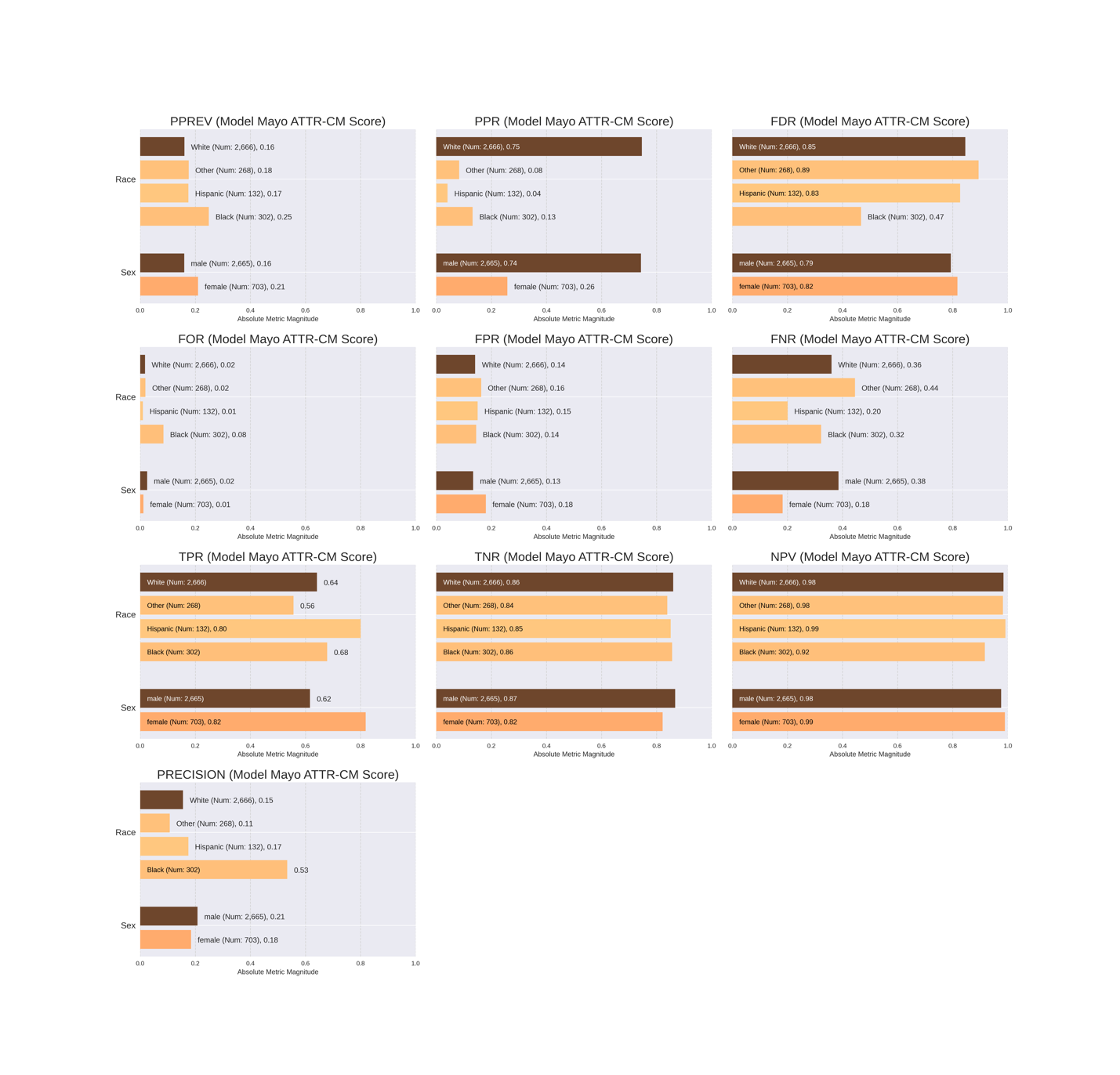

**Caption:** Darker color represents larger group size. PPREV = positive prediction rate; PPR = fraction of predicted positives belonging to each group; FDR = false discovery rate; FOR = false omission rate; FPR = false positive rate; FNR = false negative rate; TPR = true positive rate; TNR = true negative rate; NPV = negative predictive value; PRECISION = positive predictive value.

**Supplemental Figure 2. Absolute group metrics for EchoNet-LVH by race/ethnicity and sex.**

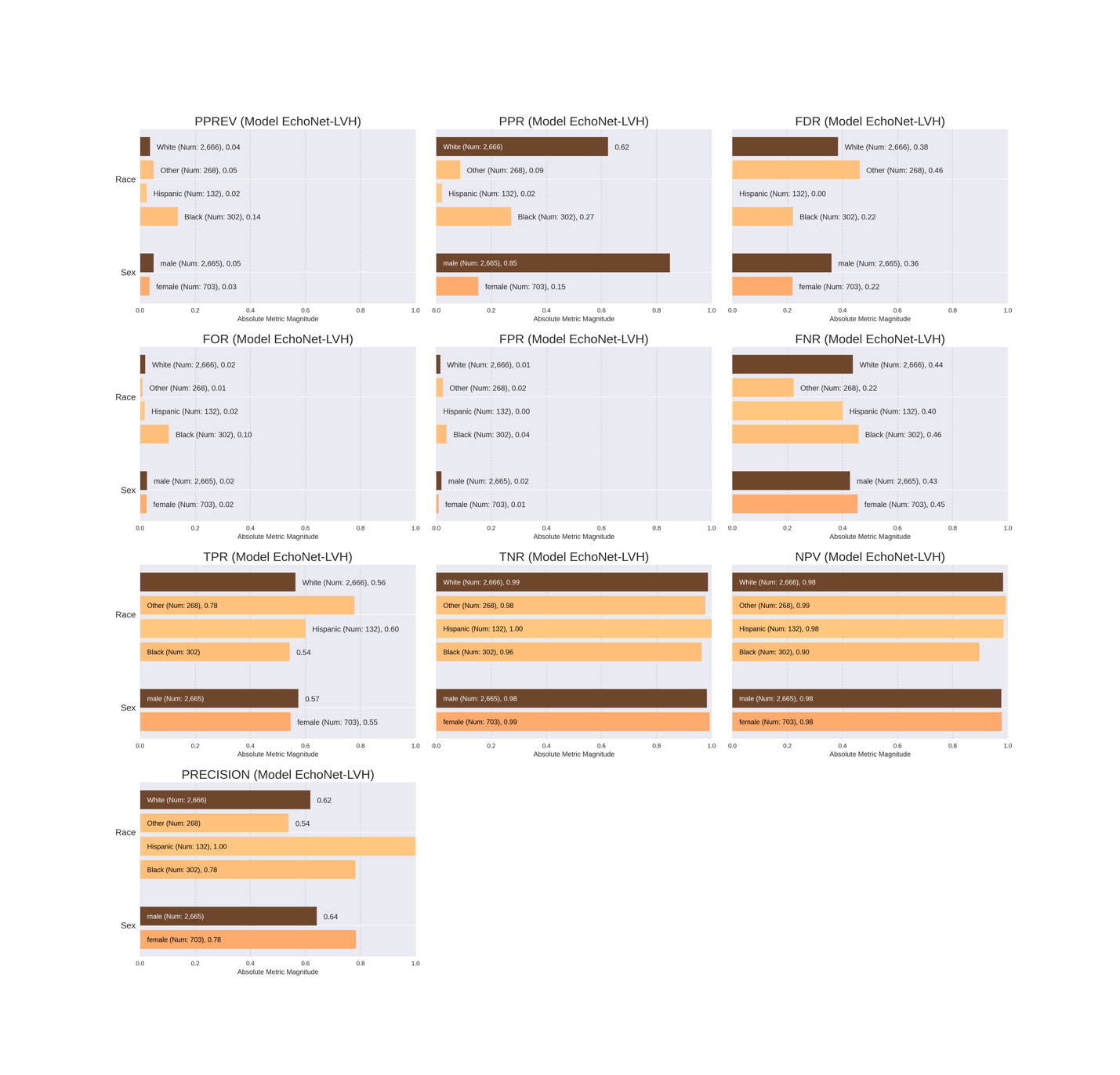

**Caption:** Darker color represents larger group size. PPREV = positive prediction rate; PPR = fraction of predicted positives belonging to each group; FDR = false discovery rate; FOR = false omission rate; FPR = false positive rate; FNR = false negative rate; TPR = true positive rate; TNR = true negative rate; NPV = negative predictive value; PRECISION = positive predictive value.

**Supplemental Figure 3. Absolute group metrics for EchoGo Amyloidosis by race/ethnicity and sex.**
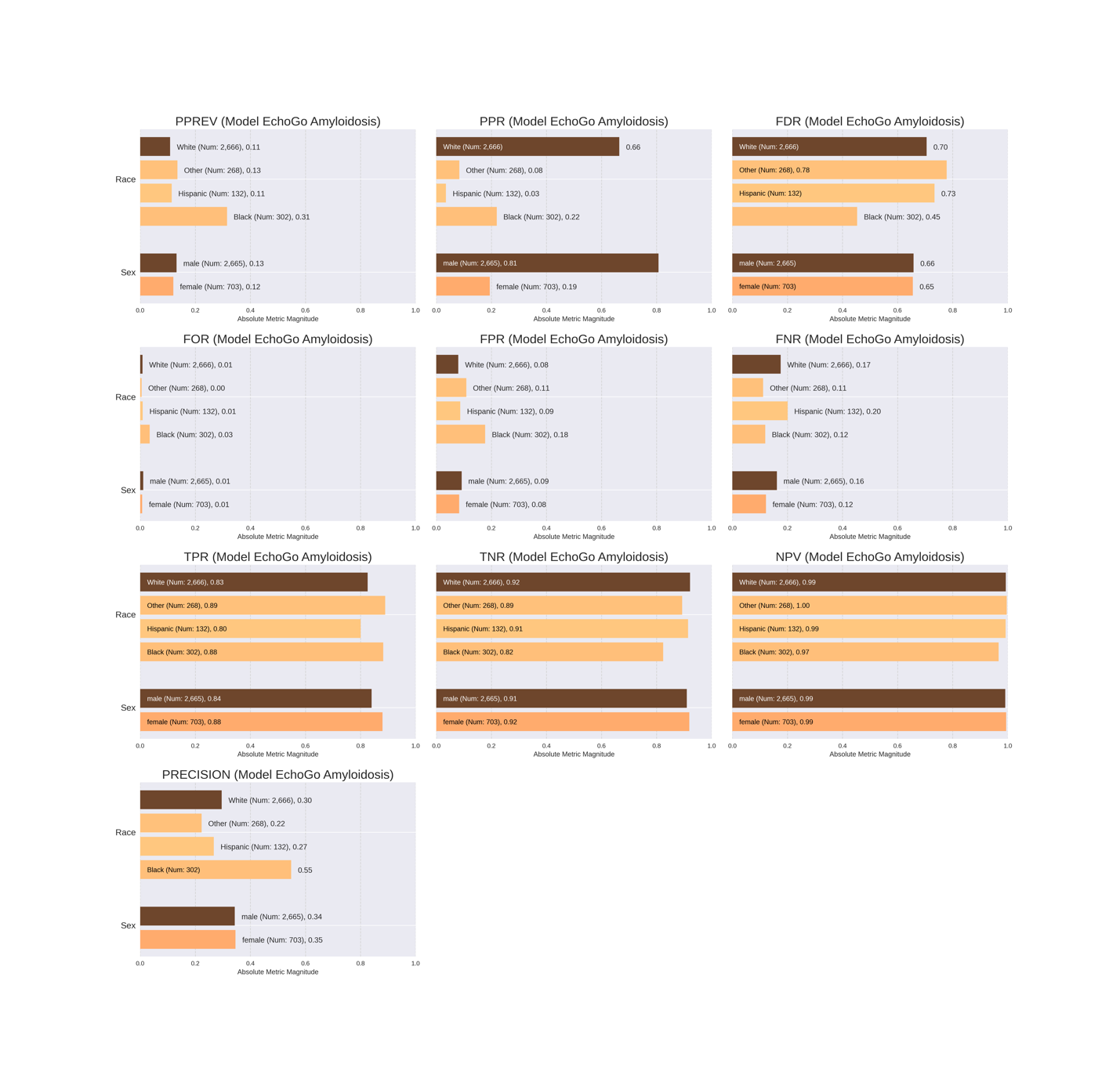

**Caption:** Darker color represents larger group size. PPREV = positive prediction rate; PPR = fraction of predicted positives belonging to each group; FDR = false discovery rate; FOR = false omission rate; FPR = false positive rate; FNR = false negative rate; TPR = true positive rate; TNR = true negative rate; NPV = negative predictive value; PRECISION = positive predictive value.

**Supplemental Table 1.** Demographic characteristics for patients with and without EchoNet-LVH predictions.

|  | **ATTR-CM** | | **HF Controls** | |
| --- | --- | --- | --- | --- |
| EchoNet-LVH Prediction | **Present (n=198)** | **Missing^a^ (n=42)** | **Present (n=3726)** | **Missing^a^ (n=834)** |
| Age, mean (SD) | 78.3 (9.2) | 75.1 (8.8) | 78.8 (9.5) | 77.3 (10.6) |
| Sex, n (%) |  |  |  |  |
| Female | 39 (19.7%) | < 10^b^ | 794 (21.3%) | 173 (20.7%) |
| Male | 159 (80.3%) | -^b^ | 2932 (78.7%) | 661 (79.3%) |
| Race/Ethnicity, n (%) |  |  |  |  |
| Black | 65 (32.8%) | 18 (42.9%) | 277 (7.4%) | 80 (9.6%) |
| Hispanic | < 10^b^ | < 10^b^ | 148 (4.0%) | 31 (3.7%) |
| White | 115 (58.1%) | 20 (47.6%) | 2981 (80.0%) | 649 (77.8%) |
| Other/Unknown | -^b^ | < 10^b^ | 320 (8.6%) | 74 (8.9%) |
| Left Ventricular Ejection Fraction , n (%) | | | | |
| < 40% | 38 (19.2%) | -^b^ | 808 (21.7%) | 208 (24.9%) |
| 40% to < 50% | 41 (20.7%) | < 10^b^ | 540 (14.5%) | 117 (14.0%) |
| ≥ 50% | 119 (60.1%) | 23 (54.8%) | 2378 (63.8%) | 509 (61.0%) |
| LVH by Posterior Wall Thickness^c^ | | | | |
| Normal/Mild, n (%) | 66 (33.3%) | 11 (26.2%) | 3247 (87.1%) | 704 (84.4%) |
| Moderate, n (%) | 65 (32.8%) | 17 (40.5%) | 407 (10.9%) | 115 (13.8%) |
| Severe, n (%) | 71 (35.9%) | 14 (33.3%) | 64 (1.7%) | 15 (1.8%) |
| LVH by Septal Wall Thickness^c^ | | | | |
| Normal/Mild, n (%) | 51 (25.8%) | < 10^b^ | 2340 (62.8%) | 425 (51.0%) |
| Moderate, n (%) | 55 (27.8%) | < 10^b^ | 451 (12.1%) | 85 (10.2%) |
| Severe, n (%) | 62 (31.3%) | 10 (23.8%) | 122 (3.3%) | 21 (2.5%) |
| Relative Wall Thickness |  |  |  |  |
| Mean (SD) | 0.75 (0.23) | 0.79 (0.29) | 0.47 (0.14) | 0.49 (0.16) |
| > 0.57 (%) | 153 (77.3%) | 34 (81.0%) | 757 (20.3%) | 200 (24.0%) |
| Hypertension, n (%) | 156 (78.8%) | 30 (71.4%) | 2971 (79.7%) | 649 (77.8%) |

^a^ Missing includes studies with an uncertain prediction

^b^ Cell sizes not reported due to small cell size

^c^ Sex-specific cutoffs were used from consensus recommendations^18^

ATTR-CM, transthyretin amyloid cardiomyopathy cases; HF, heart failure; LVEF, left ventricular ejection fraction; LVH, left ventricular hypertrophy; SD, standard deviation; IQR, interquartile range.

**Supplemental Table 2.** Demographic characteristics for patients with and without EchoGo Amyloidosis predictions.

|  | **ATTR-CM** | | **HF Controls** | |
| --- | --- | --- | --- | --- |
| EchoGo Amyloidosis | **Present (n=209)** | **Missing^a^ (n=31)** | **Present (n=3811)** | **Missing^a^ (n=749)** |
| Age, mean (SD) | 78.1 (9.0) | 75.0 (10.4) | 78.6 (9.7) | 78.2 (9.6) |
| Sex, n (%) |  |  |  |  |
| Female | 39 (18.7%) | < 10^b^ | 795 (20.9%) | 172 (23.0%) |
| Male | 170 (81.3%) | -^b^ | 3016 (79.1%) | 577 (77.0%) |
| Race/Ethnicity, n (%) |  |  |  |  |
| Black | 72 (34.4%) | 11 (35.5%) | 305 (8.0%) | 52 (6.9%) |
| Hispanic | < 10^b^ | < 10^b^ | 153 (4.0%) | 26 (3.5%) |
| White | 120 (57.4%) | 15 (48.4%) | 3035 (79.6%) | 595 (79.4%) |
| Other/Unknown | -^b^ | < 10^b^ | 318 (8.3%) | 76 (10.1%) |
| Left Ventricular Ejection Fraction , n (%) | | | | |
| < 40% | 43 (20.6%) | < 10^b^ | 832 (21.8%) | 184 (24.6%) |
| 40% to < 50% | 41 (19.6%) | < 10^b^ | 538 (14.1%) | 119 (15.9%) |
| ≥ 50% | 125 (59.8%) | 17 (54.8%) | 2441 (64.1%) | 446 (59.5%) |
| Left Ventricular Hypertrophy by Posterior Wall Thickness^c^ | | | | |
| Normal/Mild, n (%) | 63 (30.1%) | 10 (32.3%) | 3328 (87.3%) | 631 (84.2%) |
| Moderate, n (%) | 71 (34.0%) | 11 (35.5%) | 420 (11.0%) | 102 (13.6%) |
| Severe, n (%) | 75 (35.9%) | 10 (32.3%) | 63 (1.7%) | 16 (2.1%) |
| Left Ventricular Hypertrophy by Septal Wall Thickness^c^ | | | | |
| Normal/Mild, n (%) | 48 (16.6%) | 11 (35.4%) | 2339 (61.4%) | 426 (56.9%) |
| Moderate, n (%) | 54 (25.8%) | < 10^b^ | 446 (11.7%) | 90 (12.0%) |
| Severe, n (%) | 64 (30.6%) | < 10^b^ | 109 (2.9%) | 34 (4.5%) |
| Relative Wall Thickness | | | | |
| Mean (SD) | 0.76 (0.25) | 0.72 (0.23) | 0.48 (0.15) | 0.48 (0.15) |
| > 0.57 (%) | 164 (78.5%) | 23 (74.2%) | 785 (20.6%) | 172 (23.0%) |
| Hypertension, n (%) | 168 (80.4%) | 18 (58.1%) | 3042 (79.8%) | 578 (77.2%) |

^a^ Missing includes studies with an uncertain prediction

^b^ Cell sizes not reported due to small cell size

^c^ Sex-specific cutoffs were used from consensus recommendations^18^

ATTR-CM, transthyretin amyloid cardiomyopathy cases; HF, heart failure; LVEF, left ventricular ejection fraction; LVH, left ventricular hypertrophy; SD, standard deviation; IQR, interquartile range.

**Supplemental Table 3:** Huda et al. model performance on patients with 50 or more encounters in the electronic health record in 106 cases and 755 controls.

|  | **F1** | **Acc** | **Sens** | **Spec** | **PPV** | **NPV** | **FNR** | **AP (95%CI)** | | **AUC (95%CI)** |
| --- | --- | --- | --- | --- | --- | --- | --- | --- | --- | --- |
| Huda et al. | 0.21 | 0.61 | 0.43 | 0.63 | 0.14 | 0.89 | 0.57 | 0.19  [0.14, 0.25] | 0.56  [0.50, 0.62] | |

CI, confidence interval; F1, F1-score; Acc, accuracy; Sens, sensitivity; Spec, specificity; PPV, positive predictive value; NPV, negative predictive value; FNR, false negative rate; AP, average precision; AUC, area under the receiver-operating characteristic curve.

**Supplemental Table 4:** Sensitivity analysis for the EchoNet-LVH and EchoGo Amyloidosis models on 176 cases and 3192 controls at different thresholds.

| **Threshold** | **Model** | **F1** | **Acc** | **Sens** | **Spec** | **PPV** | **NPV** | **FNR** |
| --- | --- | --- | --- | --- | --- | --- | --- | --- |
| 0.06^a^ | EchoNet-LVH | 0.25 | 0.75 | 0.83 | 0.74 | 0.15 | 0.99 | 0.17 |
| 0.06^a^ | EchoGo Amyloidosis | 0.49 | 0.91 | 0.85 | 0.91 | 0.34 | 0.99 | 0.15 |
| 0.5^b^ | EchoNet-LVH | 0.54 | 0.94 | 0.64 | 0.96 | 0.47 | 0.98 | 0.36 |
| 0.5^b^ | EchoGo Amyloidosis | 0.55 | 0.93 | 0.81 | 0.94 | 0.42 | 0.99 | 0.19 |
| 0.8^c^ | EchoNet-LVH | 0.61 | 0.96 | 0.57 | 0.98 | 0.66 | 0.98 | 0.43 |
| 0.8^c^ | EchoGo Amyloidosis | 0.64 | 0.96 | 0.65 | 0.98 | 0.64 | 0.98 | 0.35 |

^a^ Default threshold for EchoGo Amyloidosis.

^b^ Commonly-used default threshold for machine learning decision boundaries.
^c^ Default threshold for EchoNet-LVH

F1, F1-score; Acc, accuracy; Sens, sensitivity; Spec, specificity; PPV, positive predictive value; NPV, negative predictive value; FNR, false negative rate.

**Supplemental Table 5:** Overall performance metrics with EchoNet-LVH and EchoGo Amyloidosis missing predictions imputed as 0 (negative) in 240 cases and 4560 controls.

|  | **F1** | **Acc** | **Sens** | **Spec** | **PPV** | **NPV** | **FNR** | **AP (95%CI)** | **AUC (95%CI)** |
| --- | --- | --- | --- | --- | --- | --- | --- | --- | --- |
| Mayo ATTR-CM Score | 0.28 | 0.84 | 0.63 | 0.85 | 0.18 | 0.98 | 0.37 | 0.14  [0.12, 0.17] | 0.78  [0.75, 0.81] |
| EchoNet-LVH | 0.54 | 0.96 | 0.47 | 0.99 | 0.65 | 0.97 | 0.53 | 0.51  [0.45, 0.58] | 0.76  [0.71, 0.80] |
| EchoGo Amyloidosis | 0.45 | 0.91 | 0.74 | 0.92 | 0.33 | 0.99 | 0.26 | 0.59  [0.52, 0.65] | 0.82  [0.78, 0.86] |

CI, confidence interval; F1, F1-score; Acc, accuracy; Sens, sensitivity; Spec, specificity; PPV, positive predictive value; NPV, negative predictive value; FNR, false negative rate; AP, average precision; AUC, area under the receiver-operating characteristic curve.
